# Antimalarial Pharmacotherapy Gaps in Nigerian Children Under Five: A Predictive Machine Learning Analysis of Care-Seeking, Testing, and ACT Treatment Using NDHS 2023–24

**DOI:** 10.64898/2026.09.10.26362761

**Authors:** Eloghosa Aisosa Nosa-Ihaza, Godsent Shepherd-Moses, Ebenezer Daberechi Okenwa, Uyioghosa Nosayise Nosa-Ihaza

## Abstract

**Background:** Nigeria’s malaria burden is the highest in the world, both in terms of incidence and fatalities. Case management uses the WHO’s Test, Treat, Track (T3) approach, where failures can occur at three stages (care-seeking, diagnostic testing, and treatment appropriateness), which are often combined into a single coverage measure. In this study, we split the cascade into two outcomes and compared four predictive modeling approaches to identify determinants.

**Methods:** From the Children’s Recode file of the 2023–24 Nigeria DHS, we identified 3,962 children under 5 years of age who reported having a fever in the two weeks before the survey. Gap A (access) was defined as: seeking care from any source and receiving a diagnostic blood test. Gap B (quality) was limited to the 1,776 children who had received any antimalarial, and specifically defined as receipt of an artemisinin-based combination therapy (ACT) only. To predict both outcomes, we tested and compared survey-weighted logistic regression, elastic-net regression, random forest, and gradient boosting (XGBoost) using a single held-out test partition and the area under the receiver operating characteristic curve (AUC) for all four models across both outcomes.

**Results:** The proportion of febrile children with combined access threshold was 16.0%, and 60.2% of children treated with antimalarials received an ACT. The Gap A logistic model did not reach overall statistical significance (p = 0.189); the Gap B model did (p = 0.003), with caregiver-reported financial barriers to care significantly associated with lower odds of ACT receipt (β = −0.97, p = 0.013), a finding that replicated across three of the four modeling approaches. AUCs were between 0.48 and 0.62 for the eight models, and all 95% CIs contained 0.50, meaning that none of the models could be confidently ruled out as being no better at discrimination than chance. For both outcomes, gradient boosting had the lowest AUC of the four methods, similar to what has been previously reported for a comparable treatment-cascade outcome in Mexico’s ENSANUT survey.

**Conclusion:** Limited variation in Nigeria’s malaria care-seeking, testing, and treatment cascade can be explained by household and maternal characteristics captured in standard survey data. The consistent association between financial access barriers and reduced likelihood of ACT-specific treatment suggests point-of-sale cost, rather than caregiver knowledge alone, as a target for intervention within Nigeria’s pharmacy and patent/proprietary medicine vendor distribution channels.

## 1. Introduction

Nigeria is one of the countries that have the highest malaria burden in the world, and malaria is one of the major causes of morbidity and mortality in children in sub-Saharan Africa. Nigeria reported 24.3% of the global malaria cases of 282 million and 30.3% of the 610,000 malaria deaths in 2024 (World Health Organization, 2025). The World Health Organization’s Test, Treat, Track (T3) Initiative describes the three steps in pediatric malaria services: seeking care on behalf of the child; confirming diagnosis with microscopy or rapid diagnostic testing (RDT) rather than presumptive treatment; and treating with an artemisinin-based combination therapy (ACT) if the diagnosis is confirmed (World Health Organization T3 Initiative, 2012). The three failure points — non-care-seeking, non-testing, and inappropriate treatment — are separate policy problems with separate policy solutions, but are often treated as one category of “coverage” in surveillance reporting, masking the exact location of the system’s failures.

The differences between them are reflected in Nigeria’s 2023/24 Demographic and Health Survey. The study defines its primary access outcome as children under five who reported having a fever in the preceding two weeks, accessed care from any source, and had a diagnostic blood test. Only 16.0% of children under five with a reported fever in the two weeks before the survey went to a diagnostic blood test and also sought care from any source. Overall, a proportion of children (60.2%) who received any antimalarial medicine for their fever received the WHO-recommended first-line medicine (ACT) instead of older or inappropriate medicines (WHO 2023). These two figures do not move together, and a comparable study in Northern Nigeria 10 years ago found the testing bottleneck was even more pronounced than the care-seeking bottleneck—of all febrile children, only 9.8% tested, while 76.7% received treatment of all kinds (Millar et al., 2014). Community pharmacies and patent and proprietary medicine vendors (PPMVs), the first point of contact for between 46 and 55% of Nigerian caregivers seeking care for a febrile child, are structurally well placed to expand access to care but not to expand diagnostic testing: a study of rural PPMVs found that only 1.9% reported testing a child under five before treating them for suspected malaria. This distinction between access and quality of care is directly applicable to pharmacy and community health policy, not just academic discussion.

This study proposes a framework of two stages of the diagnostic process of the T3 cascade, namely access gap (whether the febrile child ever enters the pathway of care, as captured in this analysis as care sought and tested) and quality gap (whether the treatment received is adequate given that the child has sought care and been tested, as captured here as in line with WHO guidance). To test both gaps rather than a single regression specification at each stage, we tested the same set of household, maternal, and access-related predictors against both gaps; we tested four analytic methods of increasing flexibility and compared out-of-sample predictive performance across all four. The NDHS is also well suited for this approach because it has an intact stratification structure that makes design-based Wald hypothesis testing feasible for every model used in this analysis, while modern machine-learning libraries support native observation weighting for random forest and gradient-boosting classifiers, thus enabling the use of the survey’s sampling weights in all but one of the four methods tested here.

The study therefore asks two related questions. First, what household, maternal, and access-related characteristics predict whether a child with a recent fever sought care and underwent a diagnostic test among Nigerian children under 5 years old—the access gap? Secondly, which of these same characteristics do predict whether a child who was given any treatment received an ACT versus an inappropriate alternative — the quality gap? We evaluate out-of-sample discriminative performance for four analytic approaches (survey-weighted logistic regression, elastic-net regularized regression, random forests, and gradient boosting [XGBoost]) and compare them across all four using a single held-out test partition. It also has a methodological function, aside from the substantive findings; namely, a comparison of predictive methods for a treatment-cascade outcome using Mexico’s ENSANUT survey found that gradient boosting underperformed simpler regression-based approaches despite its added complexity, achieving the lowest discrimination of four methods tested in Mexico (Mendoza-Cano et al., 2025). In this study, we test whether this finding replicates in a pediatric malaria context in Nigeria, using an independent dataset and outcome domain.

We assume that there are different, rather than identical, barriers at each step of the malaria case-management cascade — that is, the predictors of not seeking care, and the predictors of not receiving the appropriate care, or treatment, once care is sought — implying that closing these two gaps requires differentiated interventions rather than a single uniform policy response.

## 2. Literature Review

### 2.1 Malaria Burden and the Case for a Cascade Approach

Nigeria has the highest burden of malaria in the world. According to the 2025 World Malaria Report from the WHO, Nigeria accounted for 24.3% of the world’s 282 million malaria cases and 30.3% of its 610,000 malaria deaths in 2024, with children under five bearing a disproportionate share of that mortality. This is despite the 2021-2025 National Malaria Strategic Plan and significant multilateral investment in malaria prevention infrastructure, and indicates that progress in malaria prevention has not been matched by progress in malaria management after a child becomes febrile – the problem this study addresses (Federal Ministry of Health Nigeria, 2020).

This study’s outcome variables are grounded in the WHO’s Test, Treat, Track (T3) initiative, which was launched in 2012, and has set the policy framework: every case suspected of having malaria should be tested before treatment; every confirmed malaria case should receive a quality-assured antimalarial; and treated cases tracked through surveillance systems (World Health Organization T3 Initiative, 2012). Measuring access to care (as seeking care and testing) in this study, rather than access to care alone, follows directly from the T3 framework, where measuring access to care alone would underestimate the true failure of access.

### 2.2 Prior Nigeria-Specific Evidence on the Care-Seeking-Testing-Treatment Pathway

The most comparable previous work was Millar et al. (2014) in Sokoto and Bauchi States in Northern Nigeria, which also assessed the same three-stage pathway as this study. That study revealed that 76.7% of febrile children went for some treatment, but only 9.8% had a diagnostic blood test and 7.2% had a prompt ACT — with only 1.0% having the complete WHO/NMCP-recommended pathway (care sought, tested, and appropriately treated) (Babalola et al., 2020; Millar et al., 2014). Compared with this study’s finding that 16.0% of febrile children nationally both sought care and were tested, this shows encouraging improvement; however, testing coverage is not only a Northern Nigeria problem, as it is nationally (Shittu et al., 2026).

The findings of Anyasodor et al. (2023) for Nigeria DHS 2018 showed that 62.1% of caregivers postponed treatment for their children’s fever for over 24 hours, and that delay was observed among poorer and unemployed mothers and children less than 12-months (Anyasodor et al., 2023). Obasohan et al. (2021) applied a multilevel mixed-effects logistic model to NDHS data on malaria among children 6–59 months and found that both individual- and community-level factors matter for malaria outcomes in Nigeria — lending some support to this study’s own consideration (ultimately not pursued) of state-level clustering. Smaller facility- and community-based studies from Imo State and Ondo State also support the present study’s set of predictors for care-seeking variation, which are not simply knowledge-based.

### 2.3 The Role of Patent and Proprietary Medicine Vendors (PPMVs)

Approximately 60% of national health care is delivered by the private sector, which accounts for 90% of antimalarial distribution in Nigeria; about 46% of Nigerians with malaria symptoms seek care first from chemists or PPMVs before visiting a formal health care facility (Oladepo et al., 2019). Approximately 200,000 PPMVs across the country provide the initial point of contact for almost 55% of under-fives’ childhood illness episodes (Prach et al., 2015). For the current study’s definition of Gap A (access), this topic matters because most PPMVs cannot access diagnostics before treatment; one study estimated that only 1.9% of rural PPMVs said they had a point-of-care diagnostic tool before treating a child under five. Concurrently, substantial gaps in PPMV knowledge and stocking of recommended pediatric treatments were observed in Kogi and Kwara States by Treleaven et al. (2015), and more recently, in Kaduna and Lagos, the studies examined PPMV and community pharmacy level caregiver acceptance of test and treat (Akomolafe et al., 2024).

### 2.4 Machine Learning Applications to DHS-Based Treatment-Seeking Outcomes

Using multiple machine-learning algorithms on DHS treatment-seeking outcomes is a growing, though nascent, strategy in the sub-Saharan African literature. Yehuala et al. (2024) used XGBoost, random forest, decision tree, logistic regression, and Naïve Bayes on pooled sub-Saharan African DHS data, which had a high level of granularity, with an AUC of 0.958 for random forest, which was the best result among all models — a domain-adjacent precedent for this study, which uses pooled multi-country data rather than pooled sub-Saharan African DHS data. Fenta et al. (2024) similarly used ten ML classification approaches on pooled sub-Saharan African DHS data to predict acute respiratory infection, again finding tree-based methods competitive with simpler approaches. A study of pooled DHS data from 11 East African countries also examined how well six ML classifiers performed compared to traditional regression, and compared it with the level of interpretability provided by SHAP values, in a way that explicitly viewed the comparison of ML to traditional regression as a methodological contribution — as does this study (Kebede et al., 2026).

Of particular interest to this study is the study by Mendoza-Cano et al., 2025, whose analysis of uncontrolled hypertension compared the performance of logistic regression with LASSO, random forest, and XGBoost, all on the same data, and showed that random forest had the strongest discrimination (AUC 0.75), whereas XGBoost had the weakest (AUC 0.54) with XGBoost the most complex of the four algorithms tested. This finding — that more flexible models are not necessarily superior in out-of-sample discrimination when applied to moderately sized survey subsamples — motivated this study’s inclusion of XGBoost as a fourth comparator, to test whether the same pattern holds in a different country, outcome domain, and dataset.

### 2.5 Gaps This Study Addresses

In summary, there is a well-established literature on the testing gap in Nigeria’s malaria care-seeking pathway (Millar et al., 2014); there is a literature that shows that the existing structure of PPMVs is not well equipped to close the testing gap; and, in adjacent fields such as care-seeking for childhood ARI, ITN uptake, or hypertension, there is a growing literature of machine-learning comparisons of treatment-cascade predictors, although this is not to the authors’ knowledge as applied to Nigeria’s malaria pathway specifically. This study is filling that gap.

## 3. Methods

### 3.1 Data Source and Study Population

The study is based on the Children’s Recode (KR) from the National Population Commission and Federal Ministry of Health and Social Welfare (FMoHSSW) 2023-24 Demographic and Health Survey (NDHS) in Nigeria, and technically assisted by ICF and the DHS Program across 1,400 primary sampling units and 42,000 households in all 36 states and the Federal Capital Territory. The KR file includes 27,783 births of children born in the five years prior to the survey. As with the survey’s published case-management tables, the analytic sample was limited to children reported as having a fever in the two weeks before the survey interview (n = 3,962). The study followed ICF Institutional Review Board protocols, with informed consent from all respondents, and no additional IRB approval was required because the data are de-identified secondary data from the DHS Program data-request process.

### 3.2 Outcome Variables

We created two outcomes to represent different steps of the WHO Test, Treat, Track cascade.

Gap A (access): coded 1 if the child’s caregiver sought advice or treatment from any source (health facilities, health providers, and pharmacies), and the child had a diagnostic blood test (finger- or heel-prick) for malaria; coded 0 if either of these were not done. A caregiver’s decision to seek care was taken as a binary outcome of the survey item “no treatment or advice sought” as opposed to the more specific “taken to a medical facility” item because the latter is more restrictive and excludes care sought in pharmacies, which is the most common route for caregivers of people in Nigeria to seek first care (Literature Review). 4 children who responded “don’t know” to the testing item were coded missing rather than imputed. By this definition, 15.98% of febrile children had both conditions.

Gap B (quality): limited to 1,776 children (44.8% of the children with fever) who were treated with any type of antimalarial drugs. Within this sub-group, the outcome was defined as 1 for receiving an artemisinin-based combination therapy (ACT) and 0 for a child receiving a non-ACT antimalarial. This denominator (antimalarial-takers) was selected to align with the denominator used in the survey’s own published national indicator to enable direct comparison with the survey’s own published national estimate of ACT receipt among children who receive antimalarial treatment (∼57%), with a slight difference from one study to the other in the denominator, which can be attributed to the differences between the raw and design-weighted proportions. An initial design suggested using ACT receipt within 24 hours of fever onset as the quality threshold; this was not implemented because: (1) the timing variables were not part of the core DHS-8 questionnaire and were flagged as such in the DHS Recode Manual, and (2) in this round’s data, the timing variables were too sparsely populated to reliably estimate.

### 3.3 Predictor Variables

Predictors were chosen to represent household-, maternal- and access-related areas that were identified in the malaria treatment-seeking literature: geopolitical zone (six categories), urban/rural residence, household wealth quintile, mother’s highest educational attainment, religion, child’s ethnicity, child’s age in months, child’s sex, insecticide-treated net (ITN) use the night before the survey (coded from the household’s reported net-type variable, collapsed into a binary treated-net-use indicator), and four items from the survey’s “problems in accessing health care” battery — difficulty obtaining permission to seek care, difficulty affording treatment, distance to a health facility, and reluctance to travel unaccompanied (Rutstein et al., 2004). The variable Ethnicity, which was originally recorded on over 100 country-specific categories, was collapsed into eight analytically tractable groups (Hausa/Fulani, Yoruba, Igbo, Ijaw, Kanuri, Ibibio/Efik, Tiv, and an aggregated “Other” category), which were also assigned text labels by decoding and therefore not by a numeric code, resulting in a loss of the majority of the analytic sample due to a high level of quasi-complete separation in the first model specification. This is a limitation because a prior malaria episode as reported by the caregiver was not available as a separate variable in this survey round and was not replaced; it was reported as such.

The 73.7% missingness for ITN use, however, was not item missing but due to the survey’s subsampling design to obtain a detailed net-ownership roster. It was not dropped; instead, models were fit using listwise deletion, which resulted in an effective analytic sample for each model smaller than the original 3,962 children with fever, a limitation discussed.

### 3.4 Statistical and Analytic Approach

All models account for the NDHS’s complex sample design, including the survey’s primary sampling unit, sample stratum, and normalized individual sample weight variables. Due to the fact that when the sample is subdivided into strata based on fever, several strata contained only one sampled cluster, a common problem in complex surveys when analyzing a subpopulation; the survey design was declared with the single-unit-stratum handling set to recenter each such stratum’s contribution to the overall sample mean, avoiding the non-computable standard errors that would otherwise result.

An 80/20 train-test partition (fixed seed) was created once for the entire analytic sample and applied equally to all models and outcomes, so the final comparison of the different algorithms to the held-out observations is based on the same observations for all models.

Four algorithms of increasing analytic flexibility were fit to each outcome:

1. A survey-weighted logistic regression, with the full complex-survey design (Stata’s svy: logit), as a pre-registered baseline specification.
2. Elastic-net regularized logistic regression (Stata’s lassopack/cvlasso with α = 0.5, 5-fold cross-validation for the selection of the penalty). This is a Stata implementation of the model that departs from full survey representativeness by fitting the model unweighted; package testing shows that cvlasso does not accept probability weights in this Stata implementation (see the description of other algorithms below). This is not a limitation of the dataset, but a known limitation of the lassopack implementation in Stata, so readers should use elastic-net coefficients and AUC as indications of sample structure, not as population estimates.
3. Random forest (Python scikit-learn, 500 trees, minimum 5 observations per leaf): Fitted with the normalized sample weight passed to the model natively in the form of sample_weight.
4. The Gradient boosting model (with native observation weighting) is also fit using 500 boosting rounds, max depth of 4, and a learning rate of 0.05.

In all four algorithms, models for Gap A were fit on the training-partition subset of the full fever sample, and models for Gap B were fit on the training-partition subset of children who received any antimalarial. The categorical predictors were dummy-coded for the elastic-net, random-forest, and gradient-boosting models, and used as factor variables for logistic regression.

### 3.5 Model Evaluation

For each of the 4 algorithms and 2 outcomes, we calculated the area under the receiver operating characteristic curve (AUC) for the held out 20% test partition and assessed discrimination. The two Stata-fit models had their predictions computed with the help of the stored model estimates and AUC computed with roctab, which also gives an analytic 95% confidence interval; the two Python-fit models had their predictions computed with the stored model class probabilities, and

AUC computed with scikit-learn’s roc_auc_score, and 95% confidence intervals computed with a percentile bootstrap of 2,000 resamples on the held-out test predictions. We also analyzed variable importance for the models trained in the two libraries using the native impurity- and gain-based measures for random-forest and gradient-boosting models, respectively.

### 3.6 Software

We ran the survey-weighted logistic regression, elastic-net regression, and data management in Stata 15.1 using the lassopack and moremata packages developed by community members. We trained machine learning models (random forest and gradient-boosting models) in Python (3.9) using the pandas, scikit-learn, and xgboost packages.

## 4. Results

### 4.1 Sample Characteristics

This study used 3,962 children under five years of age who reported having a fever in the 14 days before the survey in the birth-history sample of the NDHS 2023–24, which accounted for 14.3% of all children under five in the sample. Of these, 61.2% lived in rural areas, and 34.8% lived in the North West zone, 19.5% in the North East, 13.2% in the North Central, 14.3% in the South East, 12.7% in the South South, and 5.5% in South West. Household wealth was segmented into 5 quintiles: 23.7% were in the poorest quintile, 19.8% were in the poorer quintile, 21.3% in the middle quintile, 19.6% in the richer quintile, and 15.6% in the richest quintile. Of all mothers, 40.1% had no formal education, 12.6% had only Primary education, 36.8% had Secondary education, and 10.5% had higher education. Religiously, 60.1% of households were Muslim, 30.5% were Christian, 8.7% were Catholic, and 0.7% were traditionalist. Hausa/Fulani accounted for 44.6%, Igbo 15.6%, Yoruba 5.0%, Ibibio/Efik 4.3%, Kanuri 3.1%, Tiv 0.7%, and another ethnic group accounted for 24.4% of children in the collapsed ethnicity grouping.

### 4.2 Outcome Prevalence

633 (15.98%) febrile children met both Gap A criteria (seeking care from any source and diagnostic blood testing), while 83.92% (3,325/3,962) failed to meet at least one of the two Gap A criteria, and four children with a “don’t know” response to testing lacked outcome status. Of the 1,776 children (44.8%) who were reported to have received any antimalarial drug, whether from the health system or another source — and irrespective of the type — 60.2% (1,070/1,776) were reported to have received an ACT, and 39.8% (706/1,776) were reported to have received a non-ACT antimalarial drug prior to the survey.

**Figure 1.**
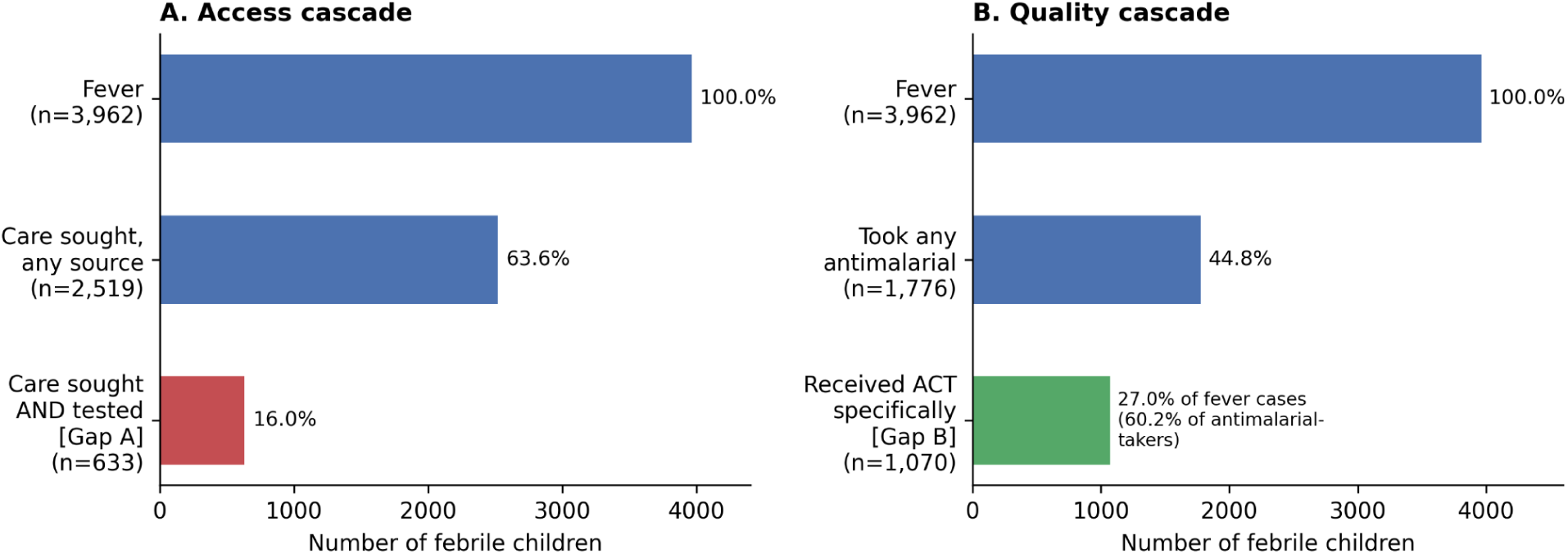
The malaria case-management cascade among children under five with fever in Nigeria is estimated from the NDHS 2023-24. *Panel A shows the access pathway (care sought, then also tested); Panel B shows the quality pathway (any antimalarial taken, then ACT specifically)*.

### 4.3 Gap A: Care-Seeking and Testing

The overall fit of the survey-weighted logistic regression on the 834 observations in the training set for Gap A, after listwise deletion, was not conventional. (F(28,363) = 1.24, p = 0.189) None of these predictors achieved the p < 0.05 level of significance; three were at the marginal significance level — rural residence (β = −0.456, p = 0.064); child’s age in months (β = 0.025, p = 0.064); and ITN used the night before (β = −1.190, p = 0.091), with the latter in an unexpected direction that should be interpreted with caution as it relates to the ITN use variable, which was missing for 73.7% of children. There was also very weak evidence of a negative association between female sex and meeting the Gap A threshold (β = −0.384, p = 0.094).

The elastic-net model included only two predictors with non-zero coefficients (λ = 95.02): being in the combined ethnicity group ‘Other’ (coefficient = 0.056; post-selection OLS coefficient = 0.126) and higher maternal education (coefficient ≈ 0.001, a nearly null effect). Because the logistic regression had a poor overall fit, this is within the scope of the few features the cross-validation could capture.

Similarly, random forest (AUC = 0.548 on 202 held-out test cases) and XGBoost (AUC = 0.479) demonstrated modest discriminative abilities for Gap A, and XGBoost failed to discriminate better than chance on this held-out partition. Both tree-based models identified child’s age as one of the strongest predictors (random forest importance = 0.249, the single largest of any variable), followed by a combination of wealth, education, zone, and access-barrier variables, which were not one major factor over the others.

**Table 1.** Gap A results across the algorithmic models.

| Gap A Model | Test AUC |
| --- | --- |
| Survey-weighted logistic regression | 0.616 |
| Elastic net | 0.545 |
| Random forest | 0.548 |
| XGBoost | 0.479 |

### 4.4 Gap B: ACT Receipt Among Antimalarial-Takers

When surveyed for antimalarials used in the 338 observations in the training set, the Gap B survey-weighted logistic regression indicated overall significant statistical findings (F(28,160) = 2.05, p = 0.003). There was a non-monotonic relationship between household wealth and receipt of ACT, with children in “poorer” (β = 0.939, p = 0.035) and “richer” (β = 1.290, p = 0.016) households having significantly higher odds of ACT than the poorest reference category, but not with “middle” and “richest” categories — a relationship that does not support a simple linear wealth gradient that is discussed further below. The Yoruba ethnic group had a higher receipt of ACT than the Hausa/Fulani group (β = 3.396, p = 0.011). Child age was significantly related to receipt of ACT: older age was associated with increased receipt (β = 0.034, p = 0.048). Perhaps most importantly, among children whose caregivers already reported a financial barrier to care, those who nonetheless obtained an antimalarial were less likely to receive the more expensive, WHO-recommended ACT specifically, rather than a cheaper alternative (β = −0.969, p = 0.013).

**Figure 2.**
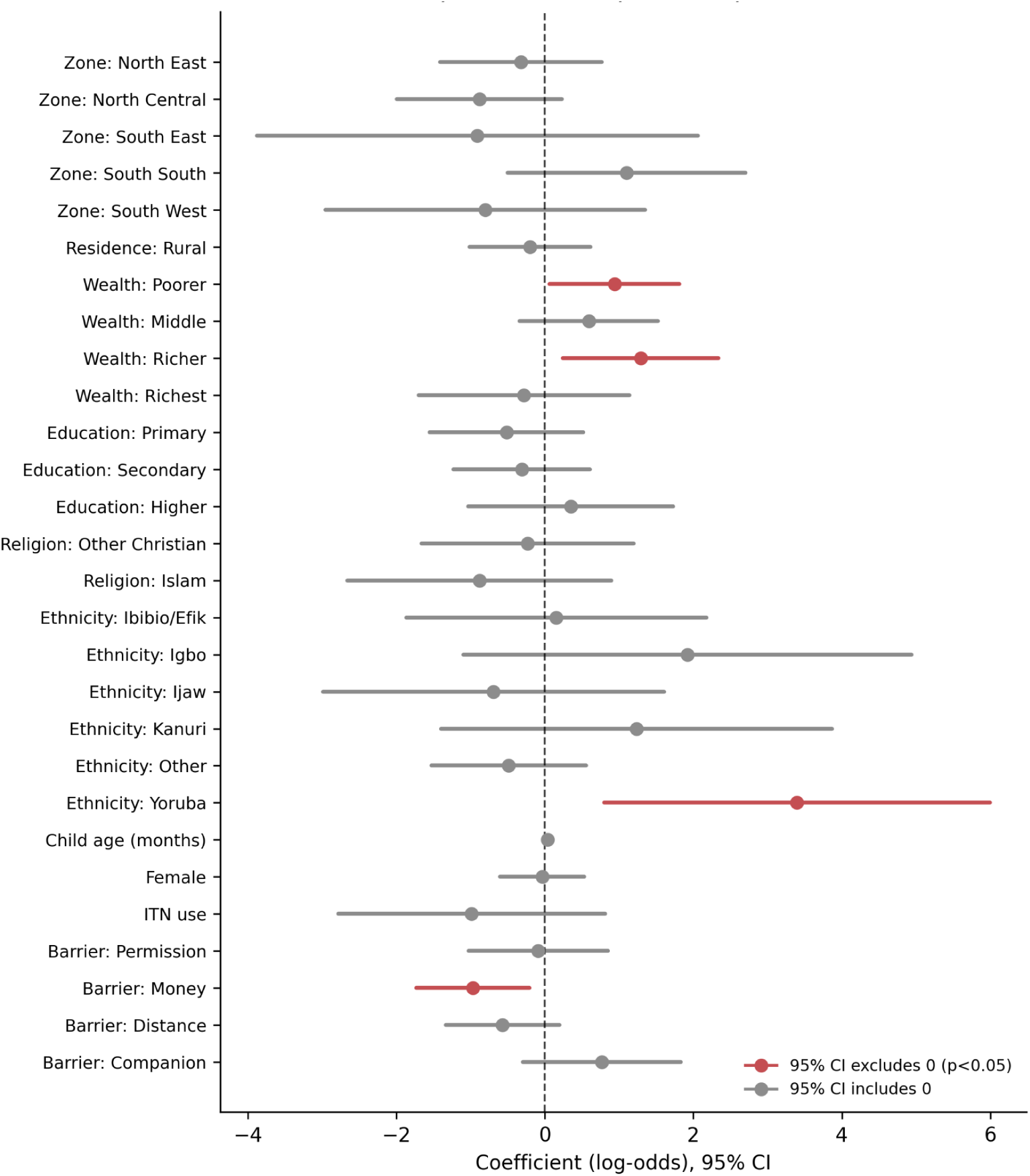
Gap B: Predictors of ACT receipt among children who were treated with antimalarials, survey-weighted logistic regression coefficients (and 95% confidence intervals). Predictors with an interval other than zero are shown by a red marker (p<0.05).

The elastic-net model retained a substantially richer predictor set than for Gap A (λ = 36.58): South South zone residence, ‘richer’ wealth quintile, Islam and traditionalist religion (both negative); five ethnicity categories (Ibibio/Efik and Igbo positive; “Other” and Tiv negative;

Yoruba strongly positive, confirming the logistic regression); child’s age and — corroborating the largest single finding from the logistic regression — the financial-access-barrier variable (coefficient = −0.036), with a negative association with reported distance to a health facility (coefficient = −0.064).

Child’s age and the financial-access-barrier were again the top features in the random forest model (AUC = 0.544, n = 71 test cases) and XGBoost model (AUC = 0.527); the Islam/religion, wealth, and ethnicity variables again featured prominently, albeit in somewhat different orders, despite the significant difference in underlying assumptions between the two models built in Stata.

**Table 2.** Gap B results across the algorithmic models.

| Gap B Model | Test AUC |
| --- | --- |
| Survey-weighted logistic regression | 0.522 |
| Elastic net | 0.589 |
| Random forest | 0.544 |
| XGBoost | 0.527 |

### 4.5 Convergence Across Models

Two findings were replicated in at least three of the four algorithms and are therefore reported with greater confidence than if they were obtained in one model alone. First, financial issues were always correlated with reduced odds of ACT-specific treatment (Gap B) in the logistic regression and in both the elastic-net and tree-based importance rankings. Second, Yoruba ethnicity was strongly associated with higher ACT receipt in both models of the logistic regression and elastic-net model for Gap B but did not appear as a leading predictor in either tree-based model in the importance analysis, and should be considered a hypothesis for further exploration because only 199 Yoruba children were included in the full sample of children with fever. Again, no single consistent pattern emerged across methods for Gap A, as this outcome was not the best throughout the rest of the analyses.

### 4.6 Comparison Across Algorithms

Across both gaps, the two methods constructed in Stata did not clearly outperform random forest, and gradient boosting performed the worst (Gap A: 0.479; Gap B: 0.527). This trend is similar to that observed in the ENSANUT Mexico comparator (Mendoza-Cano et al, 2025), which demonstrated a poor performance of gradient boosting relative to simpler regression-based treatment outcome models in a treatment-cascade treatment outcome with a sample size comparable to that of this study.

**Figure 3.**
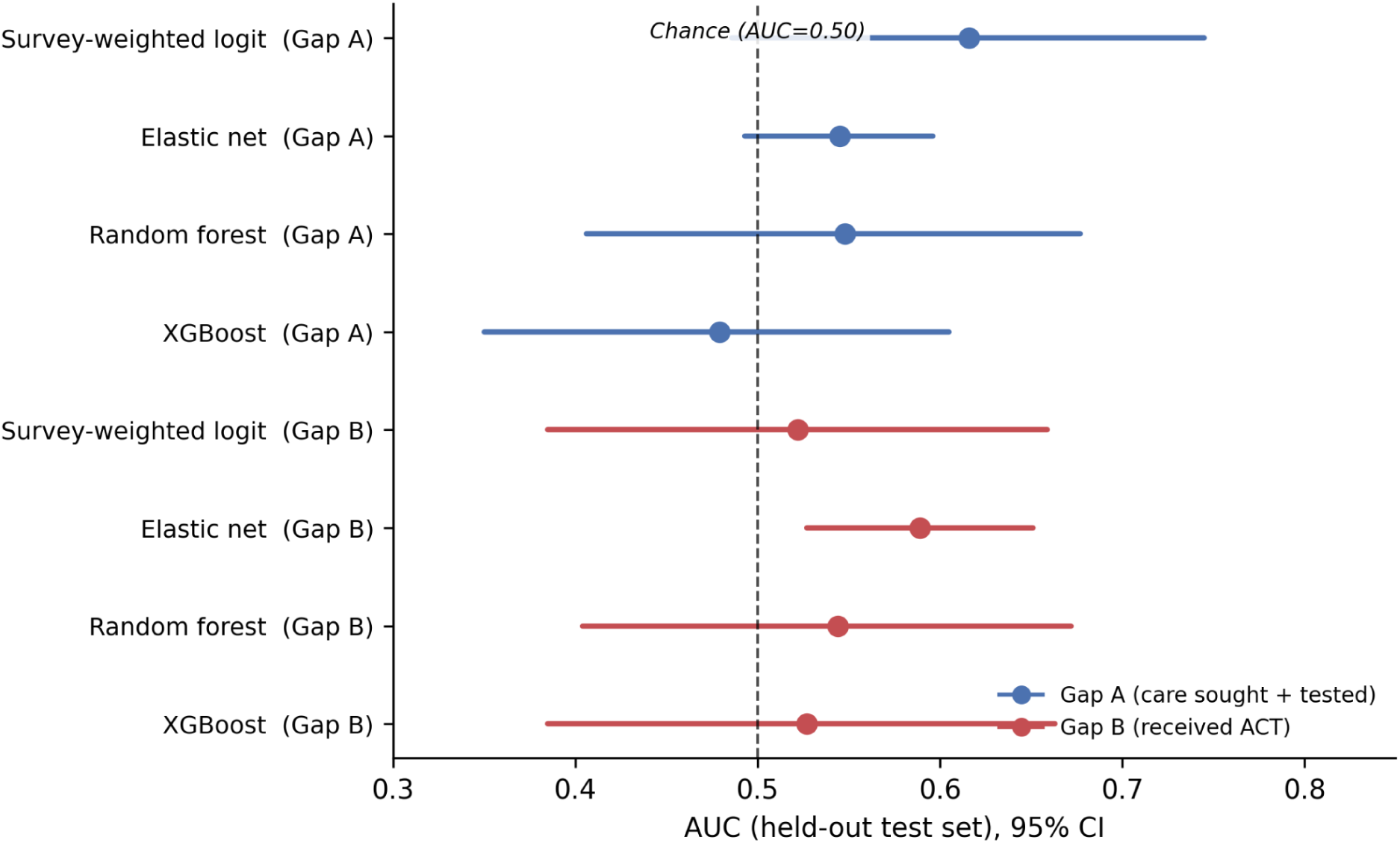
Out-of-sample discrimination (AUC) (with 95% confidence intervals) of four algorithms and both outcomes, for the held-out test set. *Each interval crosses the dashed line which indicates chance-level discrimination (AUC = 0.50)*.

## 5. Discussion

### 5.1 Principal Findings

This study aimed to empirically assess the Nigeria malaria case-management cascade for specific barriers (as stated in the Introduction), rather than uniform barriers at the access and quality stages. The findings are mixed. However, when considering individual model components, the two gaps had different significant predictors: none of the individual model components for Gap A were significant, while Gap B reached overall significance and showed a significant, interpretable relationship between financial access barriers and ACT receipt. In that very limited sense, the two gaps did different things and thus supported the hypothesis. However, the most important result was not the point estimates, but that none of the eight fitted models (four algorithms by two outcomes) could be distinguished from chance-level discrimination at 95% confidence intervals (Supplementary Table S1). The original hypothesis gets tempered a bit: It is not strictly true that different identifiable barriers exist at each stage, but rather, the household, maternal, and access characteristics of a typical DHS extract are only partly responsible for explaining who makes it to each stage, and any differences that did exist between the two gaps should be treated as suggestive and not definitive.

Nevertheless, two results are worth considering as hypothesis-generating results as they were replicated with each approach to the model more than once. First, caregiver-reported difficulty affording treatment was consistently linked to lower odds of receipt—not lower odds of receiving any antimalarial, but lower odds that those who received an antimalarial were taking the WHO-recommended, and usually more expensive, ACT. This is directly in line with the literature on Nigeria’s private medicine retail sector: these private pharmacies are patent and proprietary medicine vendors and community pharmacies, who together sell the majority of the country’s antimalarials and operate on a cash basis with greater price variation between ACTs and older, cheaper alternatives, and a caregiver already reporting a cost barrier is likely to be steered towards, or to request, the less expensive option. The finding, if it holds true in subsequent studies with bigger samples, suggests an intervention that can actually affect behavior: subsidized or capped ACT provision via the PPMV channel specifically, rather than generic messages about the superiority of ACT, which may have minimal effect on behavior when the limiting factor is the price of doing ACT rather than knowledge (Tougher et al., 2012).

Second, the relationship between household wealth and ACT receipt was not a monotonic one (Gap B) - the odds of receiving ACT were significantly higher in the “poorer” and “richer” quintiles than in the poorest quintile, but were not significantly different for the “middle” and “richest” quintiles. Most malaria-equity literature would have suggested a monotonic gradient in access to good-quality care, with the poorest households least likely to receive it, and this is what this study hoped to see. The lack of that clean gradient here doesn’t imply that wealth isn’t important; sample sizes are relatively small for individual quintiles, and a pattern can result from sampling noise in small subgroup cells. We report it rather than smoothing it into a linear interpretation for the following reasons: otherwise, we would overstate what this sample size can support.

This was also true of the replication of the ENSANUT Mexico comparator: XGBoost had the smallest AUC point estimate of four methods in both gaps, and in Gap A, the point estimate was below 0.50 throughout. Together with the precedent of ENSANUT, these two pieces of evidence come from two countries, two health domains, and two independent implementations of algorithmic flexibility, and they demonstrate that algorithmic flexibility does not reliably lead to greater out-of-sample discrimination when the underlying survey subsample is relatively small. This has a clear methodological implication for other researchers who are planning to conduct similar cascade analyses using the DHS: a four-algorithm comparison of this type is warranted because it may uncover that the more sophisticated algorithms are not providing value-added information that would not have been encountered in an analysis of one of the simpler algorithms had it been preselected.

### 5.2 Limitations

These findings are subject to several important limitations in the interpretation and application.

Small effective Analytic samples. The number of training observations and the number of test observations, as described in the models reported here, are much smaller than the base fever population, ranging from 338 to 70, depending on the model, due to listwise deletion caused by the lack of data on the insecticide-treated net (ITN) use variable for 73.7% of the fever sample. This decision was intentional: we kept the variable in the analysis rather than removing it or imputing values for households not selected in the net-roster subsample, but this is a direct result of that choice. Future analyses with access to the complete household net roster could achieve a much larger effective sample and narrower intervals, or we could accept the trade-offs of multiple imputation.

Unweighted elastic net. However, the cvlasso implementation used in the elastic-net models cannot handle probability weights, so we fit this algorithm without regard for the complex sampling design used in the survey. We confirmed this by testing the weighted syntax before reverting to the unweighted syntax, rather than relying on past precedent. The elastic-net coefficients and corresponding AUC describe this sample and should not be interpreted as design-based population estimates, as are the survey-weighted logistic regression results.

Unavailable predictor. This is because prior malaria episode, a predictor used in the original study design, was not collected as a discrete variable in a single round of any DHS, so it was omitted rather than approximated with an unvalidated proxy.

Bias due to self-report and / or self-recall. Fever status, care-seeking, testing, and treatment type are all based on a 2-week recall period from caregivers, a limitation of the DHS design shared by nearly all studies of this type. Recall reliability may be systematically related to maternal education or health literacy, which may bias the relationship between variables in either direction.

Cross-sectional design. This is a one-shot analysis, so we cannot say whether the predictors found here are stable over time or whether the small predictive power reflects a ceiling effect in this wave rather than a characteristic of the relationship. A comparison with the 2018 NDHS round, which is not included in this study, would help to differentiate these possibilities.

Ethnicity aggregation. The country-specific data from the child’s ethnicity variable (which originally had over 100 possible categories) was aggregated to eight analytically tractable groups because the data exhibited a quasi-complete separation effect in preliminary models. This was methodologically necessary to retain a usable analytic sample, but it masks potential heterogeneity within the ‘Other’ category, which accounted for 24.4% of the sample of fevers.

No cross-national comparator. As per the original study design, this analysis makes no cross-country comparison, avoiding the confound of comparing Nigeria against a country not experiencing comparable macroeconomic or health-system conditions, which is the advantage of the analysis, but it is also the disadvantage, as findings cannot be directly compared to another national context from this study alone.

### 5.3 Implications for Policy and Practice

In the context of this study’s framing of QA/pharmacovigilance, two results are worthy of attention and require a policy and pharmacy-practice perspective: First, if this financial access barrier-treatment association is replicated in larger samples, the findings indicate that the price at the point of purchase is a more direct lever for improving treatment quality than health education campaigns that target caregiver knowledge, at least for the subset of caregivers who have already surmounted the first access barrier and sought some treatment. Second, the interventions that can be implemented to address this gap are those that allow rapid diagnostic tests to be distributed in conjunction with the existing PPMVs and pharmacy distribution system, but do not necessarily need to go to formal health facilities, since the formal testing system is not the most constrained at the first point of contact (i.e., the PPMV or the pharmacy).

## 6. Conclusion

Two of the above results are worth highlighting from a policy perspective and from pharmacy practice in particular, as this study does from a QA and pharmacovigilance perspective. First, if this correlation between financial access barrier and decreased likelihood of ACT-specific treatment is replicated in larger samples, this association indicates that the financial access barrier may have a more direct impact on treatment quality than health education campaigns to increase caregiver knowledge, at least for the population of those who have surmounted the first barrier to financial access and/or visited a health care provider to begin some treatment. Second, given that only 16.0% of febrile children in this sample both sought care and received a diagnostic test — a bottleneck consistent with, though improved from, the far lower testing rates documented in earlier Northern Nigeria research — and given that PPMVs and pharmacies remain the dominant first point of contact for Nigerian caregivers but are structurally the least likely source to test before treating, interventions that pair point-of-care rapid diagnostic tests with existing PPMV and pharmacy distribution channels, rather than interventions aimed solely at formal health facilities, would target the specific bottleneck this study’s data continue to identify as the most severe.

This is not a null result to set aside. It indicates that the barriers driving non-care-seeking, non-testing, and inappropriate treatment in Nigeria are not well captured by the household and maternal characteristics typically available in cross-sectional survey data, and that supply-side factors absent from the DHS — drug stock-outs, provider-level testing capacity, out-of-pocket price at the point of sale — likely explain more of the variation than who the child is. This is the one relationship consistent across methods: a financial access barrier predicting lower odds of ACT-specific treatment. The point-of-sale mechanism this analysis identified as the clearest actionable target is the financial access barrier – interventions that reduce or subsidize the price gap between ACTs and cheaper alternatives, brought through the patent and proprietary medicine vendor channel, where most Nigerian caregivers go to obtain treatment, are more likely to close this particular quality gap than are caregiver-education campaigns.

Future research should focus on aspects not addressed by this analysis. Larger, imputation-supported samples — particularly for insecticide-treated net ownership, whose extensive missingness constrained every model here — would sharpen the confidence intervals reported in this study and clarify whether the associations observed are genuine or artifacts of limited statistical power. A repeated cross-sectional comparison against the 2018 NDHS round would help distinguish a stable predictive ceiling from a feature specific to this survey round. And supply-side data — facility stock-outs, PPMV pricing, provider training — merged with household-level DHS data would test directly whether the mechanisms this study can only infer are in fact the ones driving Nigeria’s persistent testing and treatment-quality gaps.

## Acknowledgements

The authors are thankful to Ahead Labs, IIT Roorkee, for providing Stata 15 and the necessary skillset necessary for this project. The authors also acknowledge Marwadi University for providing the resources that helped in the successful conduct of the research.

---

**Supplementary Table S1.** AUC with 95% Confidence Intervals, Held-Out Test Set.

| Outcome | Model | n(Test) | AUC | 95% CI |
| --- | --- | --- | --- | --- |
| Gap A | Survey-weighted logit | 199 | 0.616 | 0.486 – 0.745 |
| Gap A | Elastic net | 787 | 0.545 | 0.493 – 0.596 |
| Gap A | Random forest | 202 | 0.548 | 0.406 – 0.677 |
| Gap A | XGBoost | 202 | 0.479 | 0.350 – 0.605 |
| Gap B | Survey-weighted logit | 70 | 0.522 | 0.385 – 0.659 |
| Gap B | Elastic net | 335 | 0.589 | 0.527 – 0.651 |
| Gap B | Random forest | 71 | 0.544 | 0.404 – 0.672 |
| Gap B | XGBoost | 71 | 0.527 | 0.385 – 0.663 |

*Logistic regression and elastic net results computed with 95% CIs from Stata's roctab on the held out test set; random forest and XGBoost results computed with 2,000-resample percentile bootstrapping of held-out test predictions. All intervals unweighted.*

## Declarations

### Ethics Approval and Consent to Participate

The secondary data used for analysis in this study are de-identified and publicly available from the 2023–24 Nigeria Demographic and Health Survey (NDHS). The original survey team obtained informed consent from all interviewees, and the original NDHS protocol was approved by the National Health Research Ethics Committee (NHREC), Nigeria, and the ICF Institutional Review Board (IRB). This analysis relied on de-identified secondary data that is accessible via registered, authorized access to the DHS Program repository, so it was not reviewed for additional institutional review.

### Clinical Trial Registration

N/A

### Artificial Intelligence Statement

This work is not generated by a Generative Artificial Intelligence (G.A.I.) or large language model (L.L.M.) tool. The information provided is the authors’ own views and opinions.

### Data Availability Statement

Data analyzed in this study are available from the DHS Program (https://dhsprogram.com) but are not publicly available because of restrictions to protect the respondents’ confidentiality. Access is only granted with registration and approval by the DHS Program, upon reasonable request at https://dhsprogram.com/data/available-datasets.cfm. The dataset used was the Children’s Recode file of the 2023–24 Nigeria Demographic and Health Survey (NDHS). We abstracted additional literature and information from publicly available clinical trial information and World Health Organization (WHO) reports.

### Large-Language Model (LLM)

N/A

